# Apnea Test for Brain-Death Determination: A Seven-Years Retrospective Study in a French University Hospital

**DOI:** 10.1101/2025.10.14.25337990

**Authors:** Baptiste Hirsinger, Laurent Martin-Lefèvre, Karim Lakhal, Alexandre Bourdiol, Thierry Lepoivre, RN Matthieu Delahaye, Emmanuel Canet, Jean-Baptiste Lascarrou

**Affiliations:** Medecine Intensive Reanimation, Nantes University Hospital, Nantes, France; Organ Donation Department, Nantes University Hospital, Nantes, France; Department of Anesthesiology and Critical Care at Laennec Hospital, Nantes University Hospital, Nantes, France; Surgical Intensive Care Unit, Nantes University Hospital, Nantes, France; Cardiothoracic Surgery Intensive Care Unit, Nantes University Hospital, France; Medecine Intensive Reanimation, Motion-Interactions-Performance Laboratory (MIP), UR 4334, Nantes University Hospital, Nantes, France

**Keywords:** Brain death, apnea test, organ donation

## Abstract

**Background:** The apnea test is essential for the diagnosis of death by neurologic criteria but clinical data are scarce.

**Research Question:** How often and why apnea testing was not done or was aborted, how often the apnea test was completed, and the results of aborted and completed tests (positive/negative/inconclusive).

**Study Design and Methods:** This retrospective observational cohort study enrolled patients screened for death by neurologic criteria in four intensive care units at a French university hospital in 2015–2022. The main outcomes were the proportion of patients in whom the apnea test was not done due to safety concerns, the proportion of tested patients in whom the test was aborted because of adverse events, completed tests, and the results of tests. Adverse events were described. We measured arterial pH, PaO_2_, and PaCO_2_ at the start and end of each test. In patients with multiple tests, only the first was considered for the study.

**Results:** Of 371 patients screened for death by neurologic criteria, 52 (14%) did not undergo apnea testing, mainly because of respiratory or hemodynamic instability. Among all apnea tests, 94% were positive, 2% negative, and 4% inconclusive. The test was aborted in 34 (11%) patients due to adverse events, of which the most common was hypoxemia; five other severe adverse events were recorded. Test duration varied substantially even when considering only completed tests. The proportion of patients who became donors was not significantly different across the groups with completed tests, aborted tests, or no test.

**Interpretation:** Our data confirm that in most cases, the apnea test is conducted, completed and generally safe for diagnosing death by neurologic criteria. However, further research is needed to identify risk factors for adverse events during testing, contraindications to testing, and the optimal protocol for conducting the apnea test according to the characteristics of each patient.

## BACKGROUND

Brain death (BD), or death by neurological criteria (DNC), is now widely accepted as a definition of death. BD/DNC can occur in a patient with persistent cardiac output, which maintains oxygen and nutrient delivery to the organs, making them potentially suitable for transplantation.

In 1995, the American Academy of Neurology stated that BD/DNC can be diagnosed when three clinical criteria are met simultaneously: unresponsive coma, abolition of brainstem reflexes, and absence of spontaneous breathing.^1,2^ This last criterion must be verified by an apnea test, in which the patient is disconnected from the ventilator to determine whether spontaneous breathing then occurs in response to hypercapnia. The apnea test must be performed in patients with catastrophic brain injury, in the absence of confounding factors (e.g., sedatives, neuromuscular blocking agents, hypothermia, and severe hydroelectrolytic disorders) and after a period of preoxygenation. Passive oxygenation should be provided during the test to ensure sufficient oxygen delivery to the organs. The most widely accepted level of hypercapnia required to confirm the abolition of spontaneous breathing is now 60 mmHg. Adverse events have been reported during 15% to 50% of apnea tests and consist chiefly of hypotension, cardiac arrest, arrhythmias, hypoxemia, and pneumothorax.^3^

Ancillary tests can also be used to confirm BD/DNC. They include brain angiography, brain computed tomography angiography (CTA), electroencephalography (EEG), and ultrasound-based techniques. As shown in a recent studies,^4,5^ the use of ancillary tests varies across the world. In most countries, these tests are performed when confounding factors are present, the apnea test cannot be performed safely, or the results are inconclusive. However, legislation in some countries requires routine ancillary tests in addition to the apnea test to confirm brain death despite their utility has been matter of controversy ^6^. In France, the diagnosis of BD/DNC requires three steps (eFigure 1). First, BD/DNC is suspected based on the medical history and clinical findings (unresponsive coma of known origin and abolition of brainstem reflexes). Second, an apnea test is required by law without the need for informed consent ^7^ and must show absence of spontaneous breathing in response to hypercapnia. According to French guidelines, the recommended PaCO_2_ level by arterial blood gas analysis at the end of the apnea test was 50 to 60 mmHg from 2005 to 2019.^8^ However, since 2019, a level of 60 mmHg is required.^9,10^ The third step is confirmation of the diagnosis by an EEG or CTA (BD/DNC can be affirmed on the basis of step 1 and step 3 only, if the patient was judged too unstable to receive apnea test).

**Figure 1:**
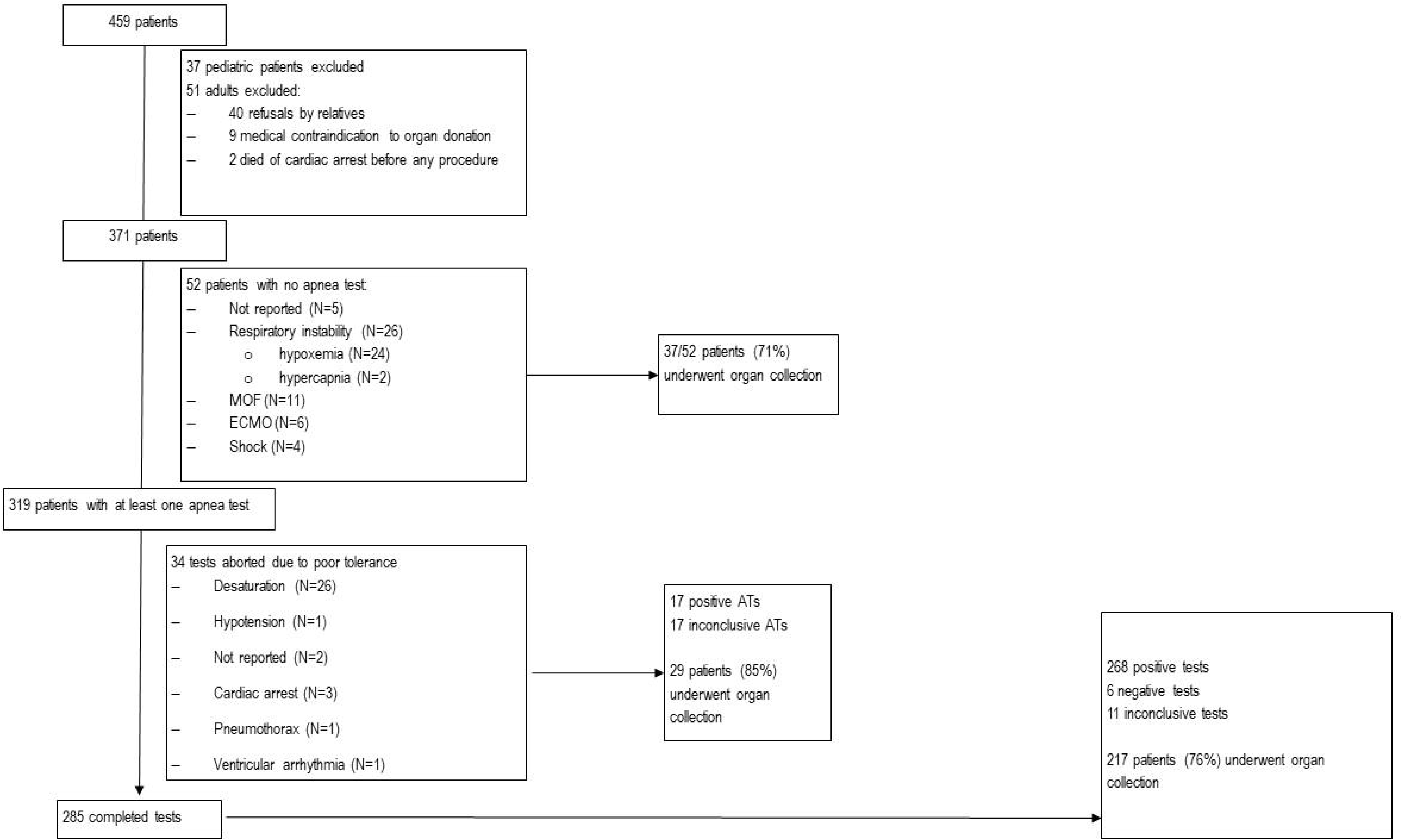
**Patient flowchart MOF: multiorgan failure; ECMO: extracorporeal membrane oxygenation**

The apnea test is traditionally performed by physically disconnecting the tracheal tube from the ventilator and providing oxygen through a cannula at a constant flow, either via a T-piece or by direct insertion into the trachea. This method can cause barotrauma and/or pneumothorax. Modifications have been suggested, such as adding a pH target to the PaCO_2_ target,^11^ limiting the test to 5 min instead of 10 min,^12^ or using continuous positive airway pressure (CPAP) for oxygenation during the test, a method reported to decrease adverse effects and improve lung-function biomarkers.^13,14^ Another modification consists in keeping the patient on the ventilator and decreasing the tidal volume and/or respiratory rate to reach the PaCO_2_ target then checking for respiratory movements.^15,16^

None of these methods has been proven optimal to affirm absence of brainstem function. In addition, organs retrieved from extended-criteria donors are being increasingly transplanted, and these donors are more likely to experience adverse events during apnea testing. In France, the mean age of donors was 38 years in 1998 and 58 years in 2023,^17^ with 43% of donors being older than 65 years. A single standardized protocol may not be optimal for all potential donors, and personalization may be desirable.

Descriptive clinical data on apnea-test practices is crucial to determine which methods ensure maximal efficacy and minimal harm to potential transplants. A single-center retrospective study from the USA described 228 apnea tests done in 1996–2007,^18^ and the same group reported 147 additional tests done in 2008–2018.^19,20^ At another center, 512 tests were done, including 484 successfully. Acidosis and a high alveolar-arterial O_2_ gradient predicted failure in this study and in another.^21,22^ A test that is aborted due to adverse events may still be valid if the PaCO_2_ level at test termination is sufficient, but the results of aborted tests are rarely reported. No published data on apnea test practices in France are available.

The objective of this retrospective cohort study was to collect data on the conduct and results of apnea tests done in several ICUs at a single center in France.

## METHODS

### Study design

We conducted a retrospective study in four intensive care units (ICUs): a medical ICU, a surgical ICU and trauma center, a mixed neurologic ICU, and a thoracic- and cardiovascular-surgery ICU. We searched the ICU databases to identify all patients in whom BD/DNC was suspected between 2015 and 2022. Exclusion criteria were age<18 y and contraindication to organ donation before an extensive neurological assessment due, for instance, to refusal of the next-of-kin or to multiorgan failure.

### Data collection

For each patient, data was extracted from the ICU electronic medical records and recorded into case report form (Excel, Microsoft, Redmond, USA). The collected data included demographics, cause of brain injury, and clinical parameters. For each apnea test, we recorded the changes in pH, PaO_2_, and PaCO_2_ during the test; test duration; and whether the test was aborted, with the reason (hypoxemia, pneumothorax, hemodynamic instability, ventricular arrhythmia, or cardiac arrest). For patients who had multiple apnea tests performed, only the first test was considered.

### Apnea Test

No specific written protocol was available to guide clinicians in performing the apnea test. The traditional method involved preoxygenating the patient for 10 minutes with an FiO□ of 1.0, then disconnecting the patient from the ventilator and administering oxygen at a rate of 3 to 5 L/min using a T-piece. Arterial blood gases were drawn at 10 minutes before reconnecting the patient to the ventilator.

### Definition of apnea test results

The recruitment period overlapped two sets of French guidelines, with a final PaCO_2_of 50–60 mmHg recommended until 2019 and a final PaCO_2_ of at least 60 mmHg subsequently. We therefore defined a positive test as PaCO_2_>50 mmHg with no respiratory movements, a negative test as PaCO_2_>50 mmHg with respiratory movements, and an inconclusive test as failure to reach PaCO_2_>50 mmHg after 10 minutes of disconnecting.

### Outcomes

The study outcomes were the proportion of patients with suspected BD/DNC but no apnea test; the proportion of patients with aborted tests due to adverse events; and, among patients with apnea tests, the proportions with positive, negative, and inconclusive results.

### Statistical analysis

The data were described as mean±SD if normally distributed and as median [interquartile range] otherwise. Continuous variables were compared using Student’s *t*-test or the Mann-Whitney test as appropriate. Categorical variables were compared using the chi-square test.

We first compared patients with vs. without an attempted apnea test. Among patients with an attempted test, we compared those in whom the test was completed (≥10 minutes) to those in whom the test was aborted.

A sensitivity analysis in which a completed test was defined as PaCO_2_>60 mmHg instead of >50 mmHg was performed.

All statistical analyses were done using STATA version 14 (StataCorp, College Station, TX). Missing data were not imputed. We did not correct the alpha risk for multiple comparisons.

### Ethics committee

The study protocol was approved by the appropriate ethics committee (French Intensive Care Society Ethics Committee 24-012). This study report complies with STROBE guidelines.^23^

## RESULTS

### Patients

Figure 1 is the patient flowchart. Of 459 identified patients, 88 were excluded due either to age <18 years (n=37) or, in adults, to refusal by relatives, contraindication to organ donation before an extensive neurological examination, or cardiac arrest. This left 371 patients for the study, including 95 (25.6%) with traumatic brain injury, 165 (44.4%) with intracranial bleeding, 32 (8.6%) with ischemic stroke, 68 (18.6%) with prolonged anoxic injury due to cardiac arrest or severe shock, and 10 (2.7%) with other causes of brain injury (hyponatremia, central nervous system infection, or complications of neurosurgery).

### Groups with vs. without an attempted apnea test

No apnea test was attempted in 52 (14%) patients. The main reasons were respiratory failure (n=26, 50.0%), multiorgan failure (n=11, 21.1%), and being on extra-corporeal membrane oxygenation (ECMO) (n=6, 11.5%). Table 1 compares the patients with vs. without an attempted apnea test.

**Table 1:**
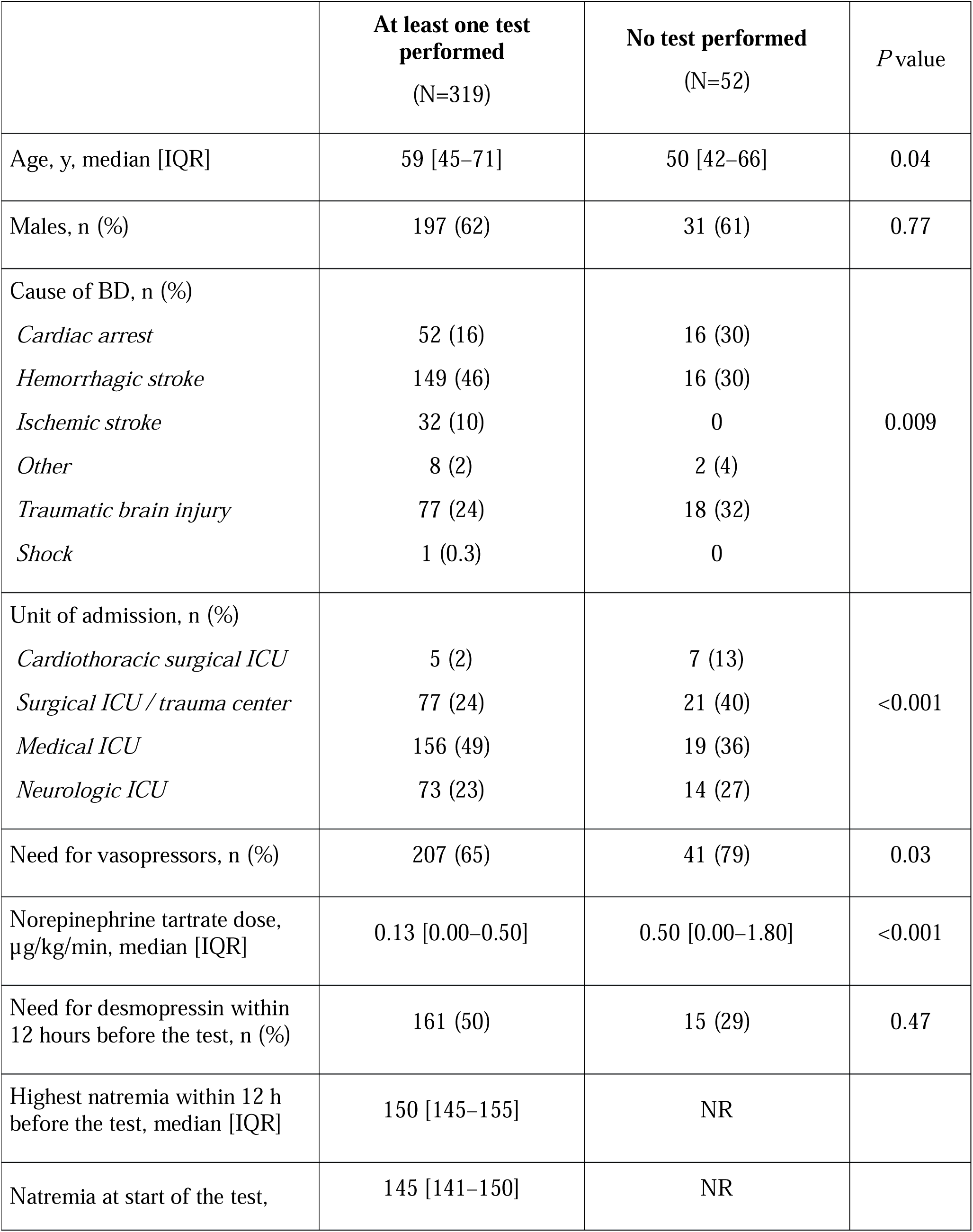
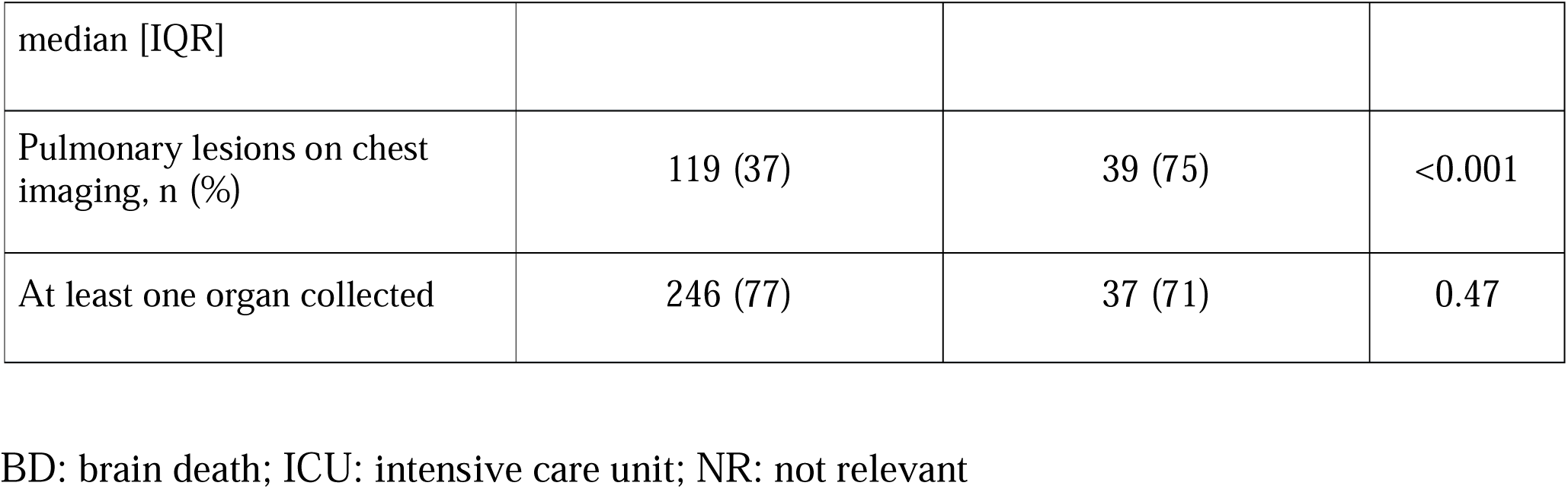
Comparison of patients with vs. without an attempted apnea test.

|  | **At least one test performed**<br>(N=319) | **No test performed**<br>(N=52) | *P* value |
| --- | --- | --- | --- |
| Age, y, median [IQR] | 59 [45–71] | 50 [42–66] | 0.04 |
| Males, n (%) | 197 (62) | 31 (61) | 0.77 |
| Cause of BD, n (%) |  |  |  |
| *Cardiac arrest* | 52 (16) | 16 (30) | 0.009 |
| *Hemorrhagic stroke* | 149 (46) | 16 (30) |  |
| *Ischemic stroke* | 32 (10) | 0 |  |
| *Other* | 8 (2) | 2 (4) |  |
| *Traumatic brain injury* | 77 (24) | 18 (32) |  |
| *Shock* | 1 (0.3) | 0 |  |
| Unit of admission, n (%) |  |  |  |
| *Cardiothoracic surgical ICU* | 5 (2) | 7 (13) | <0.001 |
| *Surgical ICU / trauma center* | 77 (24) | 21 (40) |  |
| *Medical ICU* | 156 (49) | 19 (36) |  |
| *Neurologic ICU* | 73 (23) | 14 (27) |  |
| Need for vasopressors, n (%) | 207 (65) | 41 (79) | 0.03 |
| Norepinephrine tartrate dose, $\mu\text{g/kg/min}$, median [IQR] | 0.13 [0.00–0.50] | 0.50 [0.00–1.80] | <0.001 |
| Need for desmopressin within 12 hours before the test, n (%) | 161 (50) | 15 (29) | 0.47 |
| Highest natremia within 12 h before the test, median [IQR] | 150 [145–155] | NR |  |
| Natremia at start of the test, | 145 [141–150] | NR |  |
| median [IQR] |  |  |  |
| Pulmonary lesions on chest imaging, n (%) | 119 (37) | 39 (75) | <0.001 |
| At least one organ collected | 246 (77) | 37 (71) | 0.47 |

BD: brain death; ICU: intensive care unit; NR: not relevant

**Table 2:**
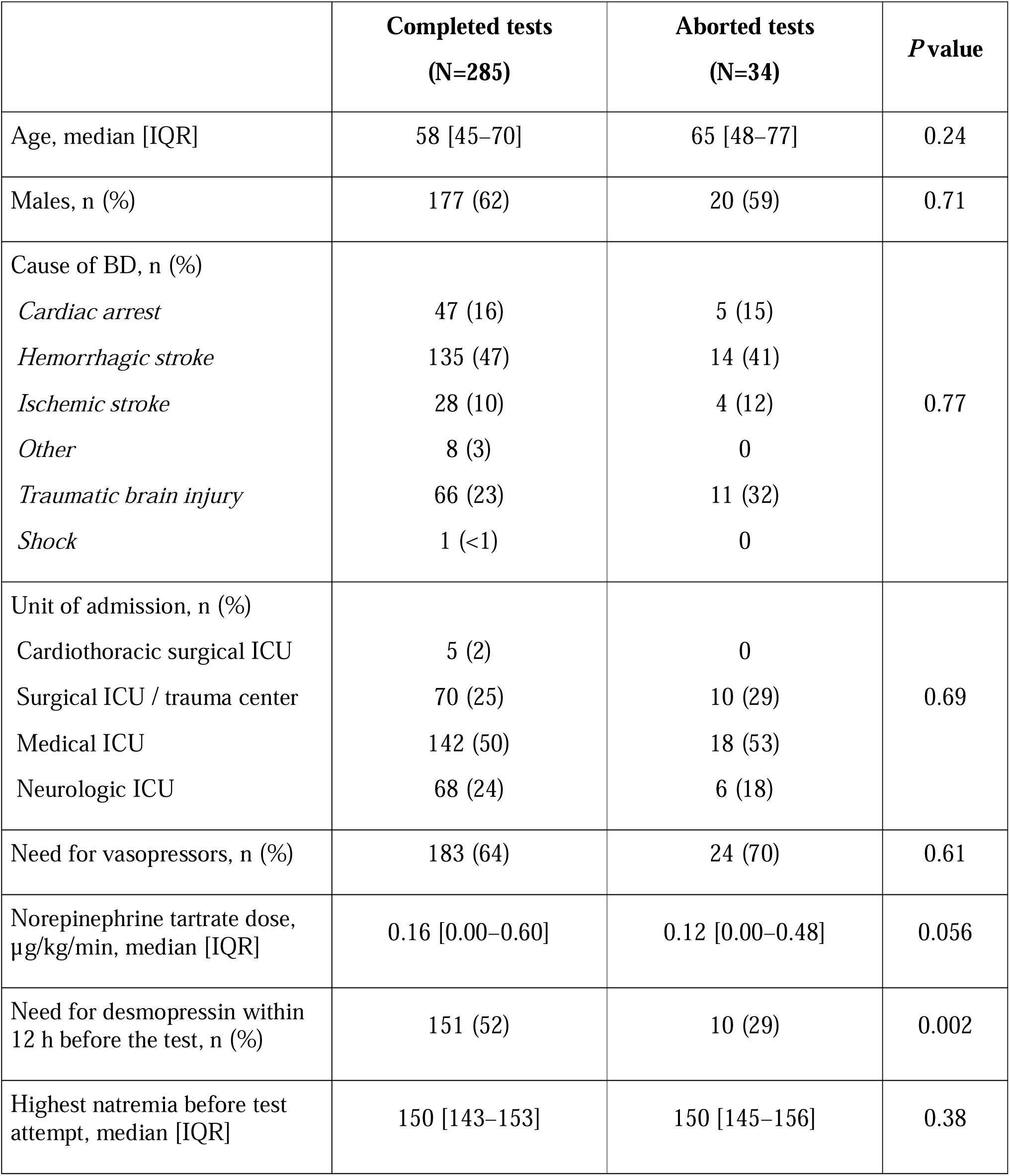
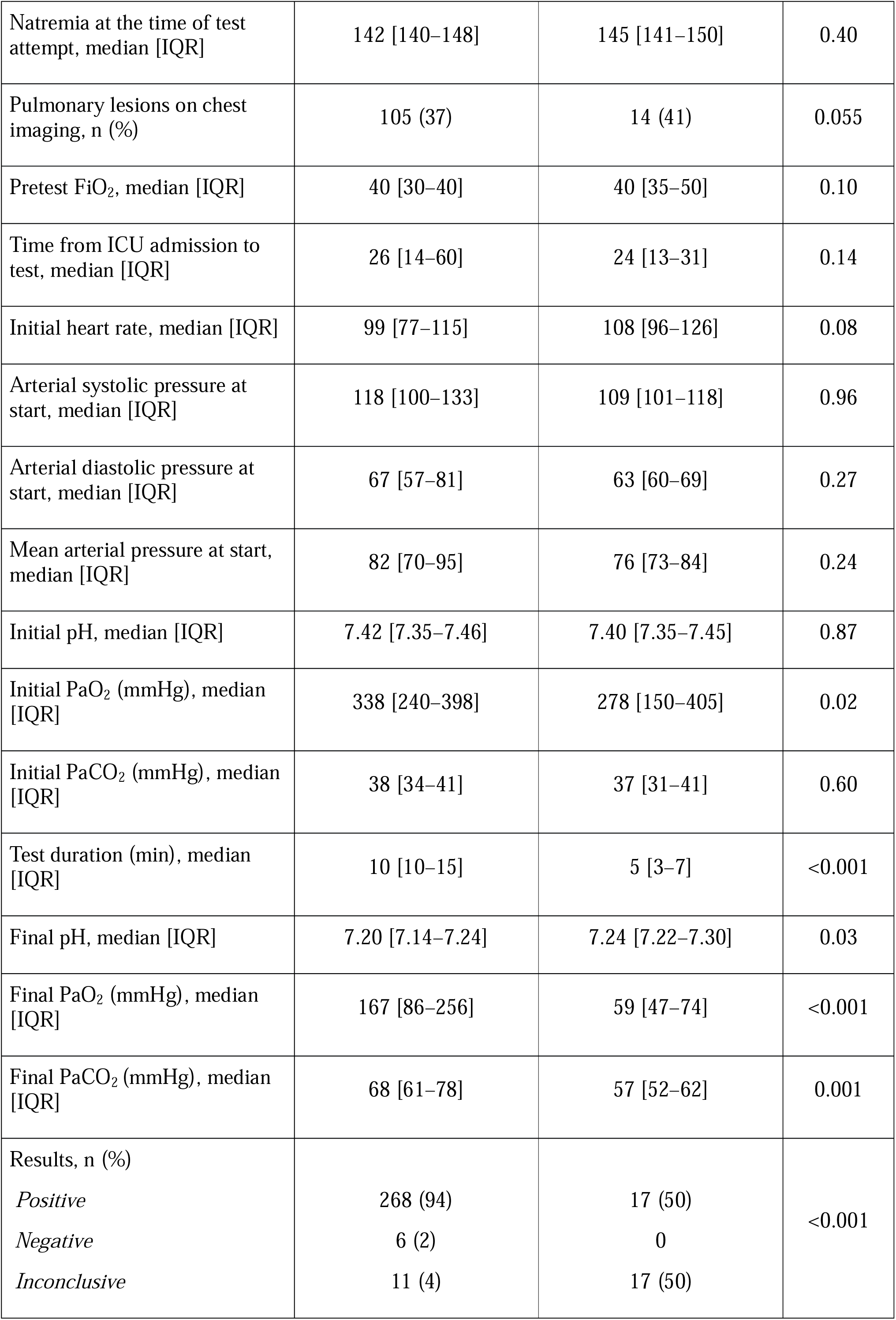
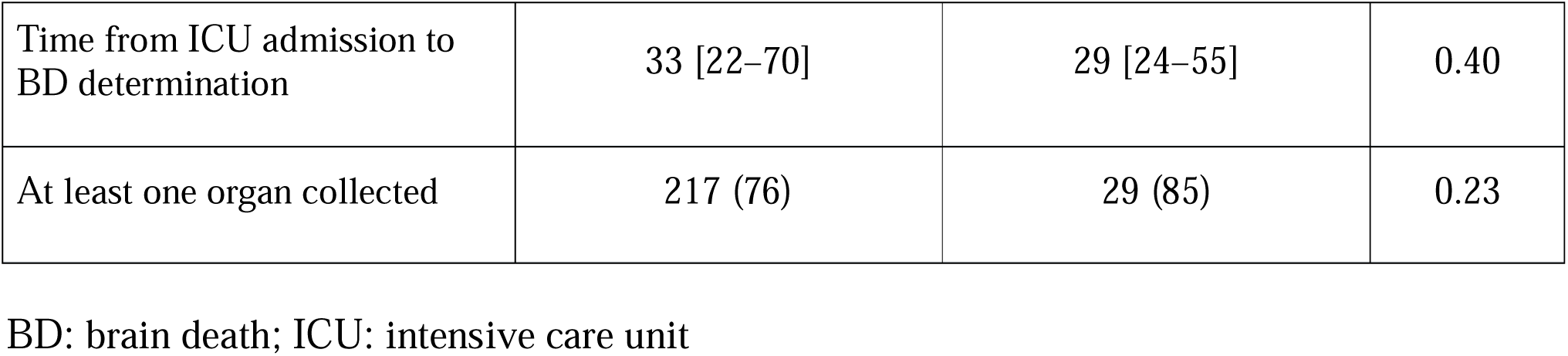
Comparison of patients completed vs. aborted apnea test.

### Groups with an aborted vs. completed apnea test

Of the 319 patients with at least one apnea test, 36 (11.3%) had the test aborted due to adverse effects including hypoxemia (n=26), cardiac arrest (n=3), hypotension (n=1), pneumothorax (n=1), and ventricular arrythmias (n=1); in two patients, the reason for aborting the test was not reported.

Aborted tests were significantly shorter than completed tests (5 min vs. 10 min, *P*<0.001). The final pH was higher (7.24 vs. 7.20, *P*=0.03), the final PaCO_2_ was lower (57 vs. 68 mmHg, *P*=0.001), and the final PaO_2_ was lower (59 vs. 167 mmHg, *P*<0.001) (Figure 2).

**Figure 2:**
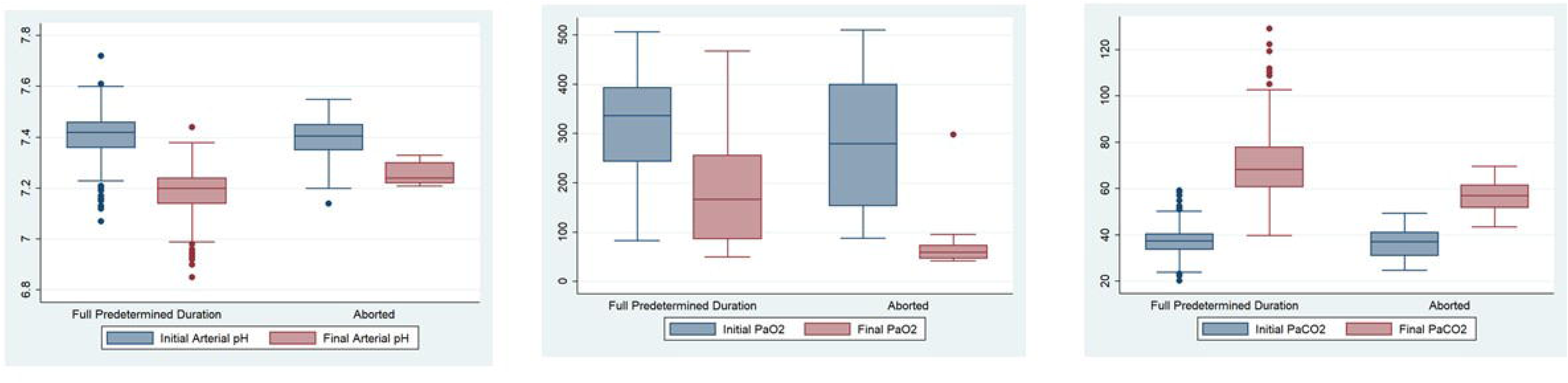
**Changes in arterial pH (a), PaO_2_ (b), and PaCO_2_ (c) during completed and aborted apnea tests**

Two pretest characteristics were associated with apnea test abortion, namely, the pretest PaO_2_ (278 vs. 338 mmHg for completed tests, *P*=0.02) and not receiving desmopressin within 12 h before the test (29% vs 52% for completed tests, *P*=0.002). Half the aborted tests could be interpreted as positive for BD. The proportion of positive tests was significantly smaller for the aborted tests (50% vs. 94% of completed tests, *P*<0.001).

Figure 2 shows the PaCO_2_ increase from the beginning to the end of the test for completed vs. aborted tests.

### Results of completed apnea tests

The median duration of the 285 apnea tests not aborted was 10 [10–15] min.

However, test duration varied substantially (Figure 3). The result was positive in 268 (94%) patients, inconclusive in 11 (4%) patients, and negative in 6 (2%) patients. Blood gas analyses showed a decrease in pH from 7.42 [7.36–7.46] at baseline to 7.20 [7.14–7.23] at completion), a decrease in PaO_2_ from 338 [240–398] mmHg at baseline to 117 [92–262] mmHg at completion, and an increase in PaCO_2_ from 38 [34–41] mmHg at baseline to 68 [62–79] mmHg at completion (Figure 2). Of the 268 positive tests, 56 ended with a final PaCO_2_ between 50 and 60 mmHg.

**Figure 3:**
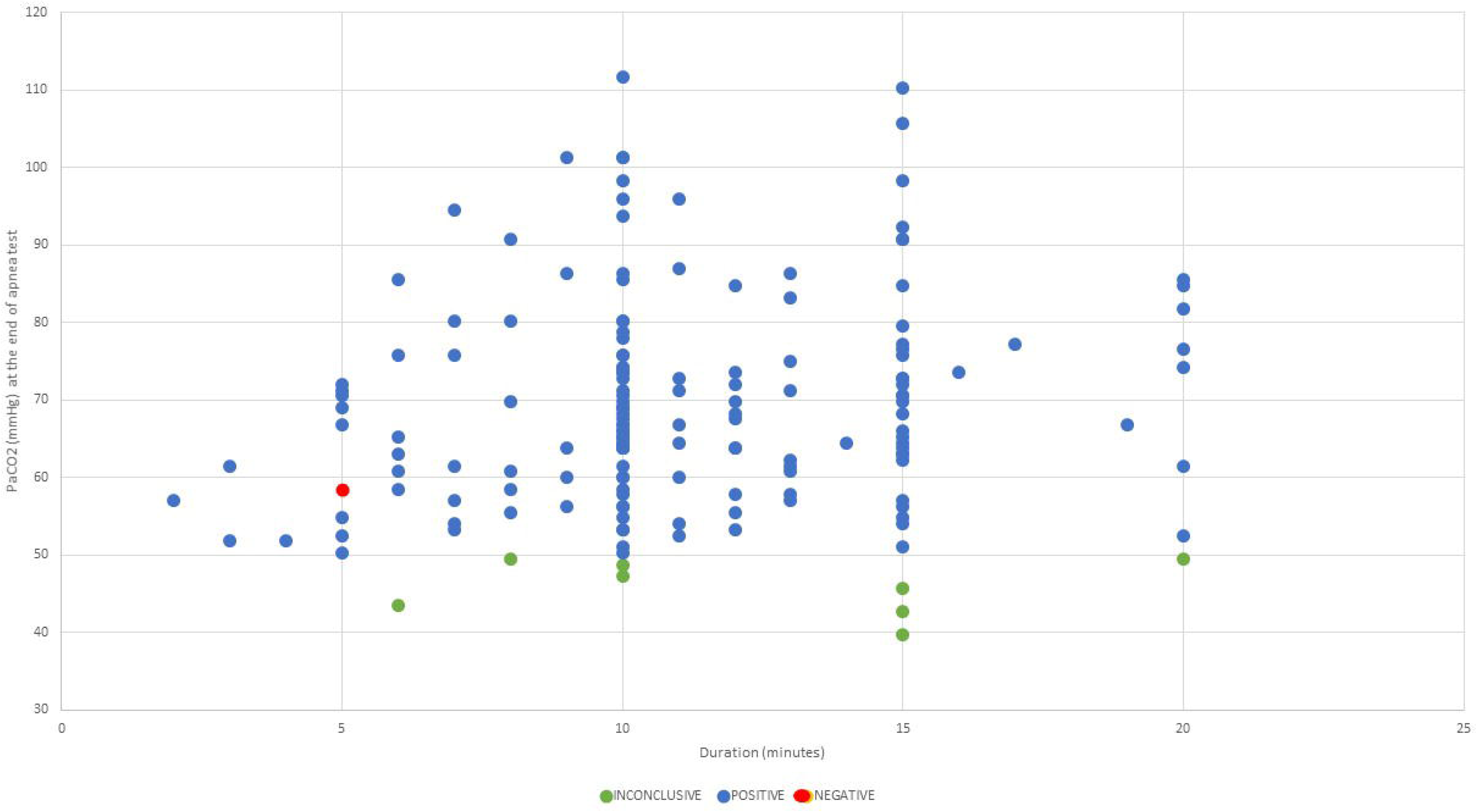
**PaCO_2_ changes according to apnea test duration**

The passive oxygenation method was reported for 101 patients. A cannula was used in 100 (99%) patients. The position of the cannula was known for only 9 patients, including 2 with the cannula in the trachea and 7 with the cannula on a T-piece. The remaining patient received CPAP with 100% FiO_2_.

Figure 3 shows the PaCO_2_ level according to test duration, and Figure 4 the proportion of positive apnea tests according to test duration.

**Figure 4:**
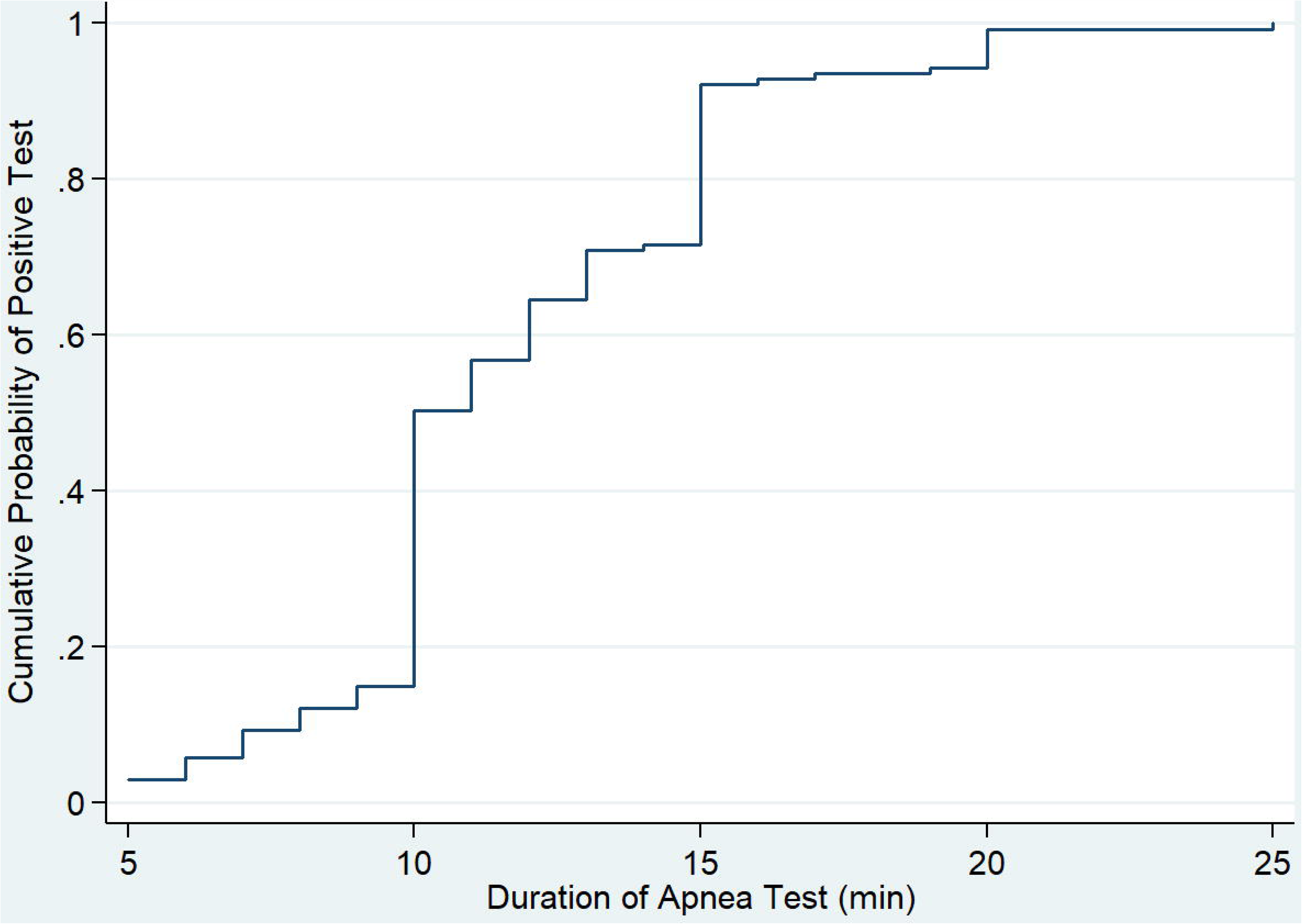
**Proportion of conclusive apnea tests according to duration**

### Negative apnea tests

In 6 patients, the test was aborted due to spontaneous breathing, which consistently occurred within the first 5 min (immediately, n=4; at 3 min, n=1; and at 5 min, n=1). Notably, the highest PaCO2 observed in those patients was 58 mmHg. All 6 patients underwent one or more subsequent apnea tests: four eventually had a positive test, one had an inconclusive test due to hypoxemia, and one had the organ donation procedure aborted at the request of the family after two negative tests (eTable 1).

### Sensitivity analysis

In the sensitivity analysis, absence of breathing with a PaCO_2_ cutoff of 60 mmHg defined a positive test. In this analysis, 56 (19.6%) completed tests and 27 (79.4%) aborted tests were considered inconclusive.

## DISCUSSION

Our retrospective cohort study found that at least one apnea test was performed in 86% of patients with suspected BD/DNC. Of the 319 patients with at least one apnea test, only 36 (11.3%) had the first test aborted due to adverse events. Of the 285 patients with a first complete apnea test, 94% (78% with a PaCO2 cut-off at 60mmHg) had a positive result, indicating BD/DNC.

Of the 371 patients eligible for the study, 47 (12.7%) were deemed too unstable to undergo apnea testing. In previous studies, the proportion of patients with no apnea test attempts ranged from 4.3%^19^ to 30%.^11^ Although respiratory or hemodynamic instability was the main reason for not performing an apnea test, the pretest FiO2 was not significantly different between patients with vs. without an apnea test. The vasopressor infusion rate was significantly higher in the untested patients but remained moderate. Conceivably, more subtle markers of instability such as rapid changes in vasopressor doses or O_2_ requirements may not have been collected for our study. In France, a positive apnea test is required by law for the diagnosis of BD/DNC (except if patient was judged too instable), and every effort is therefore made to perform the test.^24^ One option in unstable patients is to provide organ support until sufficient stability is obtained, allowing an apnea test.

Among the patients in whom an apnea test was done, 10.7% had the test aborted due to adverse events, chiefly hypoxemia. In previous studies, only 1% to 5% of patients had aborted tests. ^25^ More recently, a proportion of 11% was observed in a US cohort of 219 patients ^26^. This observation associated with our results might indicate a tendency to discontinue tests earlier nowadays, perhaps driven by the fact – in France - that all patients undergo a confirmatory ancillary test regardless of the result of the apnea test. However, our patients were older than in earlier studies (median, 48 y^21^ and 46 y^18^ vs. 59 y in our cohort). Moreover, median age in our patients with aborted tests was 65 y. Importantly, half the patients with aborted tests had positive test results confirming BD/DNC (PaCO_2_>50 mmHg without respiratory movements at the time of test termination). Among these patients with aborted tests, the proportion of donors was similar to that among patients with completed test. Thus, an arterial blood gas analysis should be performed before reconnecting the ventilator regardless of test duration or reason for abortion ^27^.

The six patients with negative apnea tests underwent at least one subsequent apnea test. A positive result was obtained in four patients and an additional patient became a donor. This finding supports repetition of the apnea test after an initial negative result. None of the six patients on ECMO had apnea tests. A review suggests that apnea tests can be performed during ECMO by reducing the sweep flow or providing CO_2_ through the ventilator.^28,29^ However, the test failure rate was high and ancillary tests were usually necessary to diagnose BD/DNC.^11^

Contrary to scientific guidelines ^10^, French law does not specify a PaCO_2_ threshold for the apnea test. In our cohort, 56 (19.6%) of the 285 completed tests ended with a PaCO_2_ between 50 and 60 mmHg, i.e. were inconclusive if using the widely accepted threshold of 60 mmHg. One patient presented spontaneous breathing at a PaCO2 of 58mmHg, supporting the use of the higher threshold of 60mmHg to ensure the absence of spontaneous ventilation.

Interestingly, PaCO_2_ at test initiation was below 33 mmHg in about a quarter of the tests. Normocapnia should be achieved before starting the test to ensure that the PaCO_2_ threshold is reached as quickly as possible.

Only 11 of the 285 completed tests were inconclusive, with failure to confirm that PaCO_2_ was >50 mmHg after ten minutes. For five patients, they had baseline hypocapnia, which should have been corrected before starting the test. In these five patients, ancillary tests confirmed BD/DNC.

The rate of PaCO_2_ increase during the apnea test may vary across patients. Point-of-care arterial blood gas assays would allow multiple PaCO_2_ measurements with test discontinuation as soon as the PaCO_2_ threshold is reached ^11^ and are therefore recommended by the 2023 American Association of Neurologists guidelines ^27^ . This would limit the risk of harm in patients whose PaCO_2_ increases rapidly. The development of transcutaneous partial CO_2_ pressure (TcpCO_2_) sensors would also allow tracking of the PaCO_2_ value during the test.

Identifying patients in whom the test will be aborted due to adverse events is challenging. In our cohort, a lower pretest PaO_2_ and not having received desmopressin were the only factors significantly associated with test abortion in our univariate analysis.

However, neither factor alone can predict test abortion. In other studies, predictors of test abortion were low systolic blood pressure, low pH, and high alveolar-arterial O_2_ gradient.^21,22^ Further studies with larger numbers of aborted tests might allow the identification of additional risk factors and, perhaps, the development of a score for predicting test abortion. Additionally, recent data about the accuracy of ancillary tests ^30^ highlights the need to secure apnea test methods in the specific context of BD/DNC.

The retrospective design is a major limitation of our study as it is associated with missing data about the apnea tests, clinical features, and laboratory results. The predefined duration of 10 minutes is questionable and could be prolonged if patient was stable during the AT. Second, all patients were recruited at the same institution, potentially limiting the generalizability of our findings to other hospitals or other countries. Strengths of our study include the large sample size and the enrollment of consecutive patients to minimize selection bias. The median age of donors (58 y) and the distribution of causes of BD/DNC (stroke, 53%; trauma, 26%, and anoxia, 21%) are consistent with recent national data on donors who died from BD/DNC.^17^

## CONCLUSION

The apnea test proved generally safe and effective for diagnosing BD/DNC in potential organ donors. However, 14% of patients were deemed too unstable to undergo apnea testing and, among tested patients, 15% had an aborted or inconclusive test. Of note, among the aborted tests, half were stopped after the PaCO_2_ threshold was reached and were positive. The frequency of severe adverse events was less than 2%. Improvements in apnea test practices would be expected to increase safety and effectiveness. More specifically, normocapnia should be achieved before starting the test and effective oxygenation methods such as CPAP should be used during the test. Tracking PaCO_2_ levels during the test by repeated blood gas sampling or by transcutaneous measurement would allow shorter test durations in patients with rapid PaCO_2_ increases. Every effort must be made to ensure that apnea testing is performed in all patients with suspected BD/DNC, as no other test can establish the diagnosis with complete certainty.

## Data sharing statement

De-identified dataset will be available upon reasonable request to corresponding author.

## Data Availability

All data produced in the present study are available upon reasonable request to the authors.

The manuscript complies with all instructions to authors from Neurocritical Care.

The authorship requirements have been met and the final manuscript was approved by all authors.

The conceptualization of the study was led by JBL. The methodology was developed by JBL. Formal analysis and data validation were carried out by BH and JBL. Validation was performed by BH and JBL, whereas investigation were managed by KL, EC, MD, AB. Data curation was done by BH and JBL, and the original draft was prepared by BH. and JBL., with review and editing by EC. All authors meet the International Committee of Medical Journal Editors criteria for authorship and have approved the final version of the manuscript.

The manuscript has not been published elsewhere and is not under consideration by another journal.

The study protocol was approved by the appropriate ethics committee (French Intensive Care Society Ethics Committee 24-012).

Conflict of Interest statement for all authors.

The authors declare no conflict of interest.

This study report complies with STROBE guidelines.

**eFigure 1: Process in our country for organ donation from donors who died of brain death**

## Notes

### Competing Interest Statement

The authors have declared no competing interest.

### Funding Statement

Not funded.

## REFERENCES

1. Practice parameters for determining brain death in adults (summary statement). The Quality Standards Subcommittee of the American Academy of Neurology. Neurology 1995;45(5):1012–1014.

2. Article R1232-2 - Code de la santé publique - Légifrance [Internet]. Available from: https://www.legifrance.gouv.fr/codes/article_lc/LEGIARTI000006909050

3. Goudreau JL, Wijdicks EFM, Emery SF. Complications during apnea testing in the determination of brain death: Predisposing factors. Neurology 2000;55(7):1045–1048.

4. Wahlster S, Wijdicks EFM, Patel PV. Brain death declaration: Practices and perceptions worldwide. Neurology 2015;84(18):1870–1879.

5. Sulmasy DP, DeCock CA, Tornatore CS, Roberts AH, Giordano J, Donovan GK. A Biophilosophical Approach to the Determination of Brain Death. CHEST 2024;165(4):959–966.

6. Wijdicks EFM. The case against confirmatory tests for determining brain death in adults. Neurology 2010;75(1):77–83.

7. Pope TM. COUNTERPOINT: Whether Informed Consent Should Be Obtained for Apnea Testing in the Determination of Death by Neurologic Criteria? No. CHEST 2022;161(5):1145–1147.

8. https://www.srlf.org/wp-content/uploads/2015/12/2005_recommandations_d_experts_prise_en_charge_des_sujets_en_etat_de_mort_encephalique_en_vue_de-prelevement_d_organes_et_de_tissus-1.pdf. [Internet] [cited 2025 June 2];Available from: https://www.srlf.org/wp-content/uploads/2015/12/2005_recommandations_d_experts_prise_en_charge_des_sujets_en_etat_de_mort_encephalique_en_vue_de-prelevement_d_organes_et_de_tissus-1.pdf

9. Gérard O. - ayant participé à la réactualisation des recommand.pdf [Internet]. 2019;Available from: https://www.agence-biomedecine.fr/IMG/pdf/prise_en_charge_des_patients_en_vue_d_un_prelevement_d_organes_-_2019_-_version_courte.pdf

10. Greer DM, Shemie SD, Lewis A. Determination of Brain Death/Death by Neurologic Criteria: The World Brain Death Project. JAMA 2020;324(11):1078–1097.

11. Migdady I, Amin M, Shoskes A. The effect of incorporating an arterial pH target during apnea test for brain death determination. J Intensive Care 2021;9(1).

12. Baik SM, Park J, Kim TY, Hong KS. Optimal duration of the apnea test for determining brain death: Benefit of the short-term apnea test. PloS One 2022;17(7).

13. Lévesque S, Lessard MR, Nicole PC. Efficacy of a T-piece system and a continuous positive airway pressure system for apnea testing in the diagnosis of brain death. Crit Care Med 2006;34(8):2213–2216.

14. Solek-Pastuszka J, Biernawska J, Iwańczuk W. Comparison of Two Apnea Test Methods, Oxygen Insufflation and Continuous Positive Airway Pressure During Diagnosis of Brain Death: Final Report. Neurocrit Care 2019;30(2):348–354.

15. Ahlawat A, Carandang R, Heard SO, Muehlschlegel S. The Modified Apnea Test During Brain Death Determination: An Alternative in Patients With Hypoxia. J Intensive Care Med 2016;31(1):66–69.

16. Kashefi P, Abbasi S, Kiani K, Khajoei MK, Akbari M. Evaluation of the new modified apnea test in confirmation of brain death. J Res Med Sci J Isfahan Univ Med Sci 2024;29(48).

17. ABM_PG_Organes_Prelevement2023.pdf [Internet]. Available from: https://rams.agence-biomedecine.fr/sites/default/files/pdf/2024-08/ABM_PG_Organes_Prelevement2023.pdf

18. Wijdicks EFM, Rabinstein AA, Manno EM, Atkinson JD. Pronouncing brain death: Contemporary practice and safety of the apnea test. Neurology 2008;71(16):1240–1244.

19. Datar S, Fugate J, Rabinstein A, Couillard P, Wijdicks EFM. Completing the apnea test: decline in complications. Neurocrit Care 2014;21(3):392–396.

20. Daneshmand A, Rabinstein AA, Wijdicks EFM. The apnea test in brain death determination using oxygen diffusion method remains safe. Neurology 2019;92(8):386– 387.

21. Kim JJ, Kim EY. Identification of Hemodynamic Risk Factors for Apnea Test Failure During Brain Death Determination. Transpl Proc 2019;51(6):1655–1660.

22. Yee AH, Mandrekar J, Rabinstein AA, Wijdicks EF. Predictors of apnea test failure during brain death determination. Neurocrit Care 2010;12(3):352–355.

23. Elm E von, Altman DG, Egger M, Pocock SJ, GÃtzsche PC, Vandenbroucke JP. The Strengthening the Reporting of Observational Studies in Epidemiology (STROBE) statement: guidelines for reporting observational studies. The Lancet 2020;370(9596):1453–1457.

24. Plourde G, Briard JN, Shemie SD, Shankar JJS, Chassé M. Flow is not perfusion, and perfusion is not function: ancillary testing for the diagnosis of brain death. Can J Anaesth J Can Anesth 2021;68(7):953–961.

25. Busl KM, Lewis A, Varelas PN. Apnea Testing for the Determination of Brain Death: A Systematic Scoping Review. Neurocrit Care 2021;34(2):608–620.

26. Kananeh MF, Brady PD, Mehta CB, et al. Factors that affect consent rate for organ donation after brain death: A 12-year registry. J Neurol Sci 2020;416:117036.

27. Greer DM, Kirschen MP, Lewis A, et al. Pediatric and Adult Brain Death/Death by Neurologic Criteria Consensus Guideline. Neurology 2023;101(24):1112–1132.

28. Giani M, Scaravilli V, Colombo SM, et al. Apnea test during brain death assessment in mechanically ventilated and ECMO patients. Intensive Care Med 2016;42(1):72–81.

29. Bein T, Müller T, Citerio G. Determination of brain death under extracorporeal life support. Intensive Care Med 2019;45(3):364–366.

30. Computed Tomography Perfusion and Angiography for Death by Neurologic Criteria | Radiology | JAMA Neurology | JAMA Network [Internet]. [cited 2025 July 1];Available from: https://jamanetwork.com/journals/jamaneurology/fullarticle/2835388

